# Chronic post-COVID neuropsychiatric symptoms persisting beyond one year from infection: a case-control study and network analysis

**DOI:** 10.1101/2023.10.22.23297069

**Authors:** Steven Wai Ho Chau, Timothy Mitchell Chue, Rachel Ngan Yin Chan, Yee Lok Lai, Paul WC Wong, Shirley Xin Li, Yaping Liu, Joey Wing Yan Chan, Paul Kay-sheung Chan, Christopher KC Lai, Thomas WH Leung, Yun Kwok Wing

## Abstract

**Background:** Limited data about chronic post-COVID neuropsychiatric complaints exist in the literature.

**Aim:** Our study aims to delineate the phenotypes of chronic neuropsychiatric symptoms among adult subjects recovering from their first COVID that occurred more than one year ago. We also aim to explore the clinical and socioeconomic risk factors of having a high loading of chronic neuropsychiatric symptoms.

**Methods:** We recruited a post-COVID group who suffered from their first pre-Omicron COVID more than a year ago, and a control group who had never had COVID. The subjects completed app-based questionnaires on demographic, socioeconomic and health status, a COVID symptoms checklist, mental and sleep health measures, and neurocognitive tests.

**Results:** The post-COVID group has a statistically significantly higher level of fatigue compared to the control group (p<0.001). Among the post-COVID group, the lack of any COVID vaccination before the first COVID and a higher level of material deprivation before the COVID pandemic predicts a higher load of chronic post-COVID neuropsychiatric symptoms. Partial correlation network analysis suggests that the chronic post-COVID neuropsychiatric symptoms can be clustered into two major (cognitive complaints -fatigue and anxiety-depression) and one minor (headache-dizziness) cluster. A higher level of material deprivation predicts a higher number of symptoms in both major clusters, but the lack of any COVID vaccination before the first COVID only predicts a higher number of symptoms in the cognitive complaints-fatigue cluster.

**Conclusions:** Our result suggests heterogeneity among chronic post-COVID neuropsychiatric symptoms, which are associated with the complex interplay of biological and socioeconomic factors.

## Background

Post-acute COVID syndrome (PACS), also known as ‘Long-COVID’, is a major global health concern (1). While SARS-CoV-2 is primarily a respiratory virus, COVID is now recognised as a multisystem disease (2) (3). Neuropsychiatric symptoms are among the most common non-respiratory symptoms in the acute phase of COVID. It is thus not surprising that some of the most commonly described symptoms of PACS are neuropsychiatric in nature, such as mood and anxiety symptoms, sleep disturbance, fatigue and cognitive complaints (4). Available evidence suggests that the risk of increased incidence in several neuropsychiatric conditions occurs not only within the first few months of SARS-CoV-2 infection, but can also extend up to a year beyond infection (5). Recent studies have also shown changes in the brain post-SARS-CoV-2 infection. A longitudinal MRI study, for example, has demonstrated structural brain changes after SARS-CoV-2 infection (6). MRI imaging studies have suggested grey matter or cortical changes among those suffering from PACS (7,8). The way SARS-CoV-2 infection affects brain health in the long term, however, is not well understood. Currently, the prevailing hypothesis links neuropsychiatric symptoms of PACS with neuroinflammation triggered by SARS-CoV-2 infection, and/or hypoxic-ischaemic insult secondary to severe respiratory distress (9).

Significant knowledge gaps also exist regarding chronic neuropsychiatric sequelae after SARS-CoV-2 infection. First, most PACS studies have focused on the first year after SARS-CoV-2 infection, leaving questions about post-COVID symptoms that run a chronic course, which is arguably a major public health concern. Second, while many PACS studies have simply considered neuropsychiatric complaints as part of a unitary PACS syndrome, this approach may overlook the heterogeneity of diverse experiences under the PACS umbrella, as well as unique factors associated with different symptom groups. For example, while preliminary evidence suggests that clinical factors, such as infection severity and COVID vaccination, are predictive factors for PACS (10), socioeconomic factors and pre-existing mental illness are often neglected in the PACS literature. Third, there have been very few studies thus far that attempt to delineate the phenotypes of chronic neuropsychiatric aspects of PACS by exploring the relationship among subjective neuropsychiatric symptoms and corresponding quantitative mental and cognitive measures.

In response to these gaps in knowledge, our current study aims to use a symptom-based approach, supplemented by validated symptoms, health-related quality of life (HRQoL) measures and app-based cognitive tests, to delineate the phenotypes of chronic neuropsychiatric symptoms among young to middle-aged adult subjects recovering from their first, pre-Omicron strain SARS-CoV-2 infection that occurred more than one year ago. We also aim to explore the relationship and clustering among common chronic neuropsychiatric symptoms, and to test if these clusters are associated with different risk factors. We set out the following hypotheses: 1. Post-COVID subjects would have a higher level of neuropsychiatric symptoms, worse neurocognitive performance and worse HRQoL one year after being first infected by SARS-CoV-2 compared to control subjects who had never been infected by SARS-CoV-2; 2. Clinical factors such as the severity of the index infection, vaccination status, and socioeconomic status will be associated with the level of chronic neuropsychiatric complaints persisting beyond one year of the index infection among post-COVID subjects; 3. Chronic neuropsychiatric symptoms among post-COVID subjects will form clusters discoverable by network analysis.

## Methods

Subjects for the post-COVID group and the control group were concurrently recruited in the community from 24 August 2022 to 1 March 2023 via posters in hospitals, online advertisements via social media, and university email newsletters. All advertising material was available in both the Chinese(in traditional characters) and English language. The inclusion criteria for the post-COVID group were as follows: (i) laboratory-confirmed SARS-CoV-2 infection which occurred before January 2022 (i.e. before the Omicron variant became dominant in Hong Kong) (11); (ii) first SARS-CoV-2 infection occurred at least one year prior to the study, and; (iii) age between 18-65 years. Inclusion criteria for the non-COVID control group were as follows: (i) no history of SARS-CoV-2 infection as confirmed by lateral-flow test or PCR; and (ii) matched with the post-COVID group in terms of the following 5 characteristics: age, gender, ethnicity, pre-COVID medical and psychiatric comorbidities, and socioeconomic status (primarily measured by the locally developed Deprivation Index, which measures material deprivation and has been widely used for local public health research (12,13)). All participants gave written informed consent either in person or via videoconference. We then instructed all participants to install a mobile phone app designed for this study. Using the app, participants completed (i) a set of questionnaires on their demographic information and socioeconomic and health status at two time points: December 2019, which was immediately before the COVID pandemic reached Hong Kong (11), and at the time of the assessment; (ii) a PACS symptoms checklist that comprised of 15 neuropsychiatric items and 26 non-neuropsychiatric items (see Supplementary materials for full checklist): the choice of items was based on results from earlier PACS studies (4), and we subsequently classified the severity of subjects’ acute COVID into 3 categories: (a) *asymptomatic or mild*, for subjects who did not have symptoms of pneumonia; (b) *moderate*, for subjects who had symptoms suggestive of pneumonia (i.e. presence of fever, cough, and shortness of breath) but did not require oxygen therapy; and (c) *severe or critical*, for subjects who had symptoms suggestive of pneumonia and required oxygen therapy and/or intensive care; (iii) a set of questionnaires on standardized mental health, sleep and HRQoL measures as follows: Patient Health Questionnaire-9 (PHQ-9) General Anxiety Disorder-7 (GAD-7), Impact of Event Scale-revised (IES-r) (14), Insomnia Severity Index (ISI) (15), Chalder Fatigue Scale (CFS) (16), CAGE screening questionnaire for alcoholism, Global Physical Activity Questionnaire (GPAQ) (17) and WHOQOL-BREF (excluding the environmental domain) (18); (iv) a seven-day sleep diary; and (v) app-based cognitive tasks, which included the psychomotor vigilance test (PVT), digital symbols substitution test (DSST), N-back test, and alternate finger tapping test (see Supplementary Method for test instructions and scoring methods). The application was available in Chinese(in traditional characters) or English depending on the subjects’ preference. Subjects who completed all assessments were compensated with HKD$150 (∼USD$20) in supermarket coupons. The authors assert that all procedures contributing to this work comply with the ethical standards of the relevant national and institutional committees on human experimentation and with the Helsinki Declaration of 1975, as revised in 2008. All procedures involving human subjects were approved the Joint Chinese University of Hong Kong-New Territories East Cluster Clinical Research Ethics Committee (Ref.: 2022.362). The study protocol was registered on the ISTCTN registry on 8 September 2022, which can be accessed at: https://doi.org/10.1186/ISRCTN35268189.

## Data Analysis

We tested for univariate between-group differences in demographics, socioeconomic and health status, and symptom score between the post-COVID group and control group using t-test, chi-square test or their nonparametric equivalent. To differentiate between post-covid subjects with a high or low symptom load, we used weighted Wasserstein distance to define a cut-off point, such that the group with a symptom number below the cut-off point (the ‘low symptom load group’) had the closest level of mental health and sleep distress, HRQoL and cognitive task performance as compared to the matched control group. The high symptom load group was then defined as the group with a symptom number above the cut-off point. To calculate the Wasserstein distance, the weights for each measurement were taken from the regression coefficients of the logistic regression model that separated the post-COVID subjects from the control group. We then performed a univariate between-group differences analysis in demographics, socioeconomic and health status, and symptom score between the high and low symptom load groups using t-test, chi-square test or their nonparametric equivalent. We used a multivariate regression model to look for potential predictors after adjusting for potential confounders.

To explore the relationships among chronic post-COVID neuropsychiatric symptoms and their clustering patterns, we built a regularized partial correlation network based on the data from the post-COVID group in relation to self-reported neuropsychiatric symptoms (19,20). To minimize the instability of the network, we excluded symptoms suffered by fewer than or equal to 20 people (out of a sample of 223 people). After network estimation, we used the *walktrap algorithm* to discover the communities/clusters within the symptom network. To validate the robustness of the detected communities, we used the *community assortativity* (r_com) metric, a bootstrapping procedure, to measure the robustness of community assignment done by the *walktrap algorithm*. Community assignments are deemed to be robust if r_com is larger than 0.5 (21). We used R packages of *bootnet*, *IsingFit, igraph, ggraph* and *plyr* to perform the network estimation process, and *asnipe* and *assortnet* for assortativity metric estimation. For details about the network estimation and validation procedures, please see Supplementary Method.

## Results

We recruited 223 post-COVID cases and 224 non-COVID control subjects (see recruitment flowchart in Supplementary Material). For the post-COVID group, the mean duration from index infection to assessment was 757 days (range = 371 – 1105 days). The majority of the post-COVID subjects had no known pre-COVID physical or mental comorbidities (80.7% and 95.5% respectively) (see Supplementary material for the full list of symptoms and their frequency), and the majority of them (70%) suffered from no or mild respiratory symptoms during the acute phase of their first COVID infection. 25% of the subjects had suffered from at least one re-infection. There were no statistically significant differences in key demographics and pre-pandemic socioeconomic and health status between the post-COVID and control groups (Table 1).

**Table 1.** List of Top Ten COVID Neuropsychiatric Symptoms (N=223)

| Symptoms | Frequency | Percentage |
| --- | --- | --- |
| Memory problems | 112 | 50.2% |
| Fatigue | 71 | 31.8% |
| Inability to concentrate | 68 | 30.5% |
| Feeling anxious | 47 | 21.1% |
| Insomnia | 46 | 20.6% |
| PTSD symptoms | 41 | 18.4% |
| Daytime Sleepiness | 39 | 17.5% |
| Feeling depressed | 34 | 15.2% |
| Headache | 30 | 13.5% |
| Loss of interest or pleasure | 29 | 13.0% |

The differences in the level of depressive, anxiety, post-traumatic stress, and insomnia symptoms between the post-COVID and control groups did not reach statistical significance. The post-COVID group, however, had significantly more fatigue symptoms than the control group (median = 3.0 vs 2.0, p<0.001) despite a similar daily sleep duration. The HRQoL measures of the two groups were comparable. In relation to app-based cognitive tasks, there was no statistically significant difference between the two groups in terms of their performance in the PVT and N-back tasks. The post-COVID group performed worse in the DSST (median of total accurate count = 58.0 vs 60.5, p = 0.013) and the alternate finger tapping task as compared to the control group (median of total accurate count = 34.0 vs 41.0, p = 0.012).

The post-COVID group had more newly diagnosed medical conditions since the beginning of the COVID pandemic (mean = 0.14 vs 0.05, p= 0.0062), but not psychiatric diagnoses (see Supplementary material for the full list of symptoms and their frequency). They also had more days of hospitalisation in the past year (median = 0 vs 0, mean = 0.9 vs 0.2, p = 0.048) but not more medical consultations. The post-COVID and control groups did not have statistically significant changes in socioeconomic status, as reflected by changes in the Deprivation Index and subjects’ employment status.

Among patients who recovered from COVID, 60% reported at least one neuropsychiatric symptom which appeared during the acute or early post-recovery period of their first COVID infection and has persisted until now. The most frequent complaints were memory problems (50.2%), fatigue (31.8%), inability to concentrate (30.5%), anxiety (21.1%), insomnia (20.6%), post-traumatic stress (PTS) (18.4%, as measured by IES-R), daytime sleepiness (17.5%) and feeling depressed (15.2%) (Table 2).

**Table 2.** Comparisons of baseline characteristics, post-pandemic changes of health and socioeconomic status, infection and vaccination status between subjects of post-COVID group and control group, and between high post-COVID neuropsychiatric symptoms load group and low symptoms load group, respectively.

| Variables | Post-COVID group | Control group | P-Value | High symptom load group | Low symptom load group | P-Value |
| --- | --- | --- | --- | --- | --- | --- |
| **Baseline Characteristics** |  |  |  |  |  |  |
| Age, year mean (SD) | 42.2 (14.4) | 41.6 (13.7) | 0.65 | 44.8 (14.0) | 41.1 (14.4) | 0.083 |
| Female Sex, % | 60.5% | 61.6% | 0.82 | 61.2% | 60.3% | 0.90 |
| Non-Chinese ethnicity, % | 2.7% | 2.7% | 0.99 | 0% | 3.8% | 0.10 |
| Tertiary education level, % | 54.7% | 62.1% | 0.12 | 49.3% | 57.1% | 0.28 |
| Pre-pandemic Deprivation Index, median (IQR) | 1.0 (3) | 0.5 (2) | 0.098 | 1.0 (4) | 1.0 (2) | 0.065 |
| Pre-pandemic no. of medical conditions, mean (SD) | 0.27(0.64) | 0.28(0.65) | 0.94 | 0.30(0.58) | 0.26(0.66) | 0.31 |
| Any psychiatric conditions pre-pandemic, % | 4.5% | 4.9% | 0.83 | 4.5% | 4.5% | 0.10 |
| Pre-pandemic suicidal ideation | 4.5% | 6.2% | 0.42 | 3.0% | 5.1% | 0.30 |
| **Post-pandemic Changes in Health and Socioeconomic Status** |  |  |  |  |  |  |
| Change in Deprivation Index, median (IQR) | 0 (0) | 0 (0) | 0.89 | 0 (0.5) | 0 (0) | 0.17 |
| No. of new medical conditions, mean (SD) | 0.14(0.42) | 0.05(0.24) | 0.0062 | 0.25(0.50) | 0.10(0.37) | 0.0022 |
| Any new psychiatric conditions, % | 2.7% | 3.1% | 0.79 | 7.5% | 0.6% | 0.004 |
| New suicidal ideation,% | 7.2% | 4.5% | 0.22 | 16.4% | 3.2% | 0.0 |
| **Infection and Vaccination Status** |  |  |  |  |  |  |
| Any COVID vaccination before infection, % | 11.2% | N/A | - | 4.5% | 14.1% | 0.037 |
| > 1 infection, % | 26.0% | N/A | - | 22.4% | 27.6% | 0.42 |
| First infection severity, % |  |  |  |  |  | 0.63 |
| Mild | 70.0% | N/A | - | 65.7% | 71.8% | - |
| Moderate | 18.8% | N/A | - | 22.4% | 17.3% | - |
| Severe or ICU admission | 11.2% | N/A | - | 11.9% | 10.9% | - |

*Note.* The high symptom load group is defined as subjects with four or more chronic neuropsychiatric symptoms, while the low symptom load group is defined as subjects with fewer than four persistent neuropsychiatric symptoms. SD: standard deviation. IQR: interquartile range.

**Table 3.**
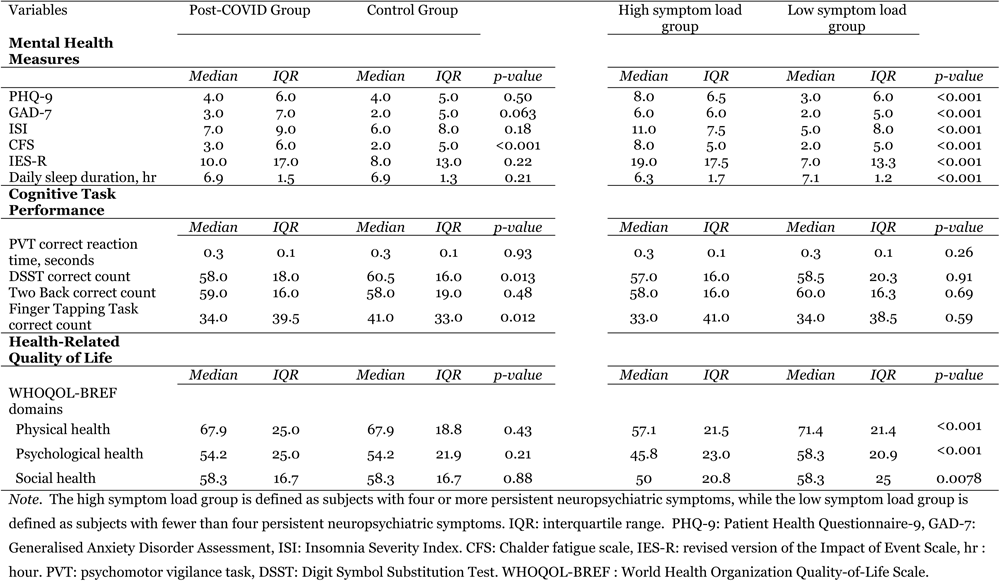
Comparison of mental health measures, cognitive task performance and health-related quality of life outcome between subjects of post-COVID group and control group, and between high post-COVID neuropsychiatric symptoms load group and low symptoms load group, respectively.

| Variables | Post-COVID Group |  | Control Group |  |  | High symptom load group |  | Low symptom load group |  |  |
| --- | --- | --- | --- | --- | --- | --- | --- | --- | --- | --- |
| **Mental Health Measures** |  |  |  |  |  |  |  |  |  |  |
|  | *Median* | *IQR* | *Median* | *IQR* | *p-value* | *Median* | *IQR* | *Median* | *IQR* | *p-value* |
| PHQ-9 | 4.0 | 6.0 | 4.0 | 5.0 | 0.50 | 8.0 | 6.5 | 3.0 | 6.0 | <0.001 |
| GAD-7 | 3.0 | 7.0 | 2.0 | 5.0 | 0.063 | 6.0 | 6.0 | 2.0 | 5.0 | <0.001 |
| ISI | 7.0 | 9.0 | 6.0 | 8.0 | 0.18 | 11.0 | 7.5 | 5.0 | 8.0 | <0.001 |
| CFS | 3.0 | 6.0 | 2.0 | 5.0 | <0.001 | 8.0 | 5.0 | 2.0 | 5.0 | <0.001 |
| IES-R | 10.0 | 17.0 | 8.0 | 13.0 | 0.22 | 19.0 | 17.5 | 7.0 | 13.3 | <0.001 |
| Daily sleep duration, hr | 6.9 | 1.5 | 6.9 | 1.3 | 0.21 | 6.3 | 1.7 | 7.1 | 1.2 | <0.001 |
| **Cognitive Task Performance** |  |  |  |  |  |  |  |  |  |  |
|  | *Median* | *IQR* | *Median* | *IQR* | *p-value* | *Median* | *IQR* | *Median* | *IQR* | *p-value* |
| PVT correct reaction time, seconds | 0.3 | 0.1 | 0.3 | 0.1 | 0.93 | 0.3 | 0.1 | 0.3 | 0.1 | 0.26 |
| DSST correct count | 58.0 | 18.0 | 60.5 | 16.0 | 0.013 | 57.0 | 16.0 | 58.5 | 20.3 | 0.91 |
| Two Back correct count | 59.0 | 16.0 | 58.0 | 19.0 | 0.48 | 58.0 | 16.0 | 60.0 | 16.3 | 0.69 |
| Finger Tapping Task correct count | 34.0 | 39.5 | 41.0 | 33.0 | 0.012 | 33.0 | 41.0 | 34.0 | 38.5 | 0.59 |
| **Health-Related Quality of Life** |  |  |  |  |  |  |  |  |  |  |
|  | *Median* | *IQR* | *Median* | *IQR* | *p-value* | *Median* | *IQR* | *Median* | *IQR* | *p-value* |
| WHOQOL-BREF domains |  |  |  |  |  |  |  |  |  |  |
| Physical health | 67.9 | 25.0 | 67.9 | 18.8 | 0.43 | 57.1 | 21.5 | 71.4 | 21.4 | <0.001 |
| Psychological health | 54.2 | 25.0 | 54.2 | 21.9 | 0.21 | 45.8 | 23.0 | 58.3 | 20.9 | <0.001 |
| Social health | 58.3 | 16.7 | 58.3 | 16.7 | 0.88 | 50 | 20.8 | 58.3 | 25 | 0.0078 |

*Note.* The high symptom load group is defined as subjects with four or more persistent neuropsychiatric symptoms, while the low symptom load group is defined as subjects with fewer than four persistent neuropsychiatric symptoms. IQR: interquartile range. PHQ-9: Patient Health Questionnaire-9, GAD-7: Generalised Anxiety Disorder Assessment, ISI: Insomnia Severity Index. CFS: Chalder fatigue scale, IES-R: revised version of the Impact of Event Scale, hr : hour. PVT: psychomotor vigilance task, DSST: Digit Symbol Substitution Test. WHOQOL-BREF : World Health Organization Quality-of-Life Scale.

### Defining symptom load groups

The levels of depression, anxiety, fatigue, insomnia, PTS, as well as HRQoL measures, but not cognitive performance, worsened as the number of chronic neuropsychiatric symptoms reported increased (see Supplementary material). The weighted Wasserstein distance analysis showed that post-COVID subjects with less than four persistent neuropsychiatric complaints were closest to the matched control group in terms of the level of mental and sleep problems, HRQoL and cognitive task performance (Figure 1). Thus, post-COVID subjects with less than four persistent symptoms were defined as the low symptom load group, while those with four symptoms or more were defined as the high symptom load group.

**Figure 1.**
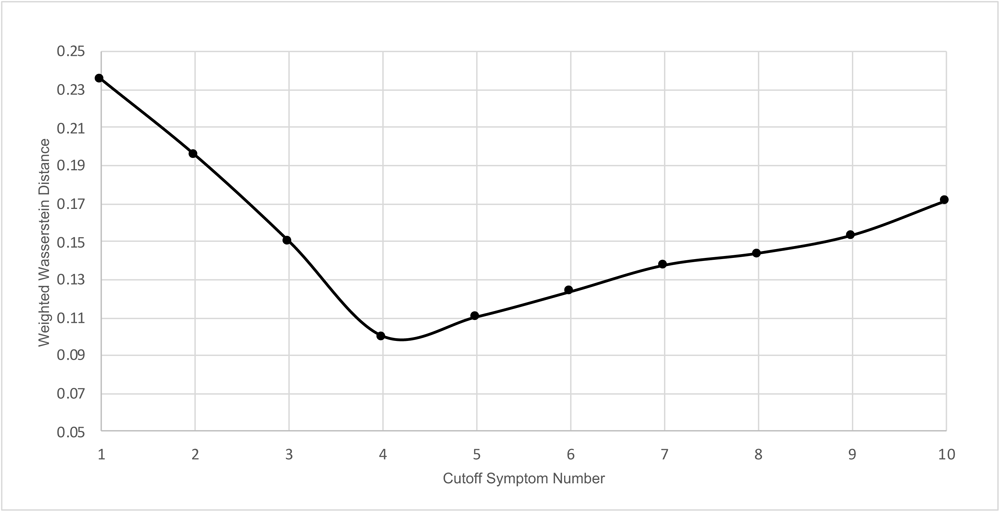
Optimal threshold for defining high/low symptom load groups.

**Figure 2.**
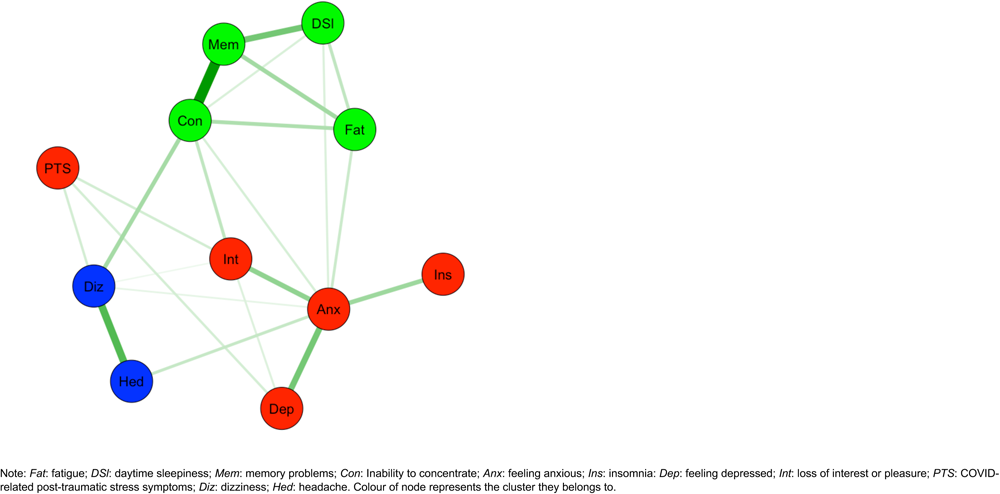
Partial correlation network of post-COVID chronic neuropsychiatric symptoms

According to this cut-off, 67 of our subjects (30%) had a high symptom load. There were no statistically significant differences in the baseline characteristics of the high and low symptom load groups, although the mean age of the high symptom load group was nominally higher (mean = 44.8 vs 41.1, p = 0.083), and the difference in pre-COVID levels of social deprivation between the groups approached statistical significance (median = 1.o vs 1.o, p=0.065). The median number of pre-morbid medical conditions and the presence of known psychiatric conditions in the two groups were similar, though the high symptom load group had a higher number of persistent non-neuropsychiatric symptoms after the acute phase of COVID infection (median = 6 vs 0, p<0.001).

The high symptom load group had a statistically significantly higher number of newly diagnosed medical conditions (median = 0 vs 0, mean = 0.3 vs 0.1, p = 0.002) and a statistically significantly higher incidence of new psychiatric conditions (7.5% vs 0.6%, p=0.004) and suicidal ideas (16.4% vs 3.2%, p<0.001). Additionally, they had more medical consultations (median = 3.0 vs 1.0, p<0.001) and days of hospitalisation (median = 0 vs 0, mean = 0.7 vs 1.0, p=0.007) in the past year.

The high symptom load group was less likely to have received any COVID vaccine prior to their first SARS-CoV-2 infection (4.5% vs 14.1%, fisher exact test p= 0.038) when compared to the low symptom load group, but the two groups were similar in terms of the proportion of moderate to severe/critical severity of the subjects’ COVID infection during the acute phase (34.3% vs 28.2%, p= 0.43) and whether they had single or multiple COVID infections (22.4% vs 27.6%, p = 0.51).

A multivariate logistic regression performed using age, gender, ethnicity, tertiary education, pre-pandemic Deprivation Index (log-transformed), number of pre-pandemic medical comorbidities, presence of pre-pandemic psychiatric diagnosis, receipt of COVID vaccination prior to first infection, severity of index COVID infection and single or multiple infections as predictors showed that only the pre-COVID Deprivation Index (adjusted odd ratio = 1.55, 95% CI = 1.04-2.31, p= 0.032) and receipt of any COVID vaccination prior to first infection (adjusted odd ratio = 0.26, 95% CI = 0.07-0.92, p= 0.037) were statistically significant predictors for the high symptom load group.

### Network analysis and symptom clustering

Our network model identified three distinct clusters/communities: (1) an anxiety-depression cluster containing symptoms of anxiety, insomnia, loss of interest or pleasure, depression and PTSD; (2) a cognitive complaint-fatigue cluster containing fatigue, inability to concentrate, memory problems and daytime sleepiness; and (3) a dizziness-headache cluster. The r_com of the network was 0.68, indicating our network model likely contained discrete clusters and that the *walktrap algorithm* had reliably detected them.

Multivariate regression models using the same predictors as our logistic regression above suggested that the pre-COVID Deprivation Index (log-transformed) positively predicts the number of chronic symptoms in both the anxiety-depression (b=0.32, p=0.013) and cognitive complaint-fatigue clusters (b=0.29, p=0.029), but receipt of any COVID vaccination before the index infection negatively predicts only the number of symptoms from the cognitive complaint-fatigue cluster (b=-0.72, p=0.023) and not that of the anxiety-depression cluster (b=-0.43, p=0.15). In addition, being ethnically Chinese predicted a higher number of symptoms from the cognitive complaint-fatigue cluster (b=-1.26, p= 0.038).

## Discussion

To the best of our knowledge, our study is the first that examines chronic neuropsychiatric symptoms persisting beyond one year after the first SARS-CoV-2 infection using (i) non-COVID control data to define a threshold for high/low symptom load and (ii) network analysis to explore the structure of such symptoms, as well as the clinical and socioeconomic risk factors of symptom clusters.

### Higher level of fatigue and worse performance in neurocognitive tasks among post-COVID subjects

The post-COVID group had a higher level of fatigue than the matched control group after more than one year from the index infection, a finding which was not explained by depression, anxiety, PTS, or insomnia. This result echoes existing evidence that fatigue is one of the most reported PACS symptoms in the post-infection era (22), with the additional implication that fatigue may be more persistent than mood, anxiety, sleep or PTS symptoms among post-COVID patients. The post-COVID group also performed worse than the matched control group in the DSST and alternate finger-tapping tests. The DSST is a nonspecific psychomotor speed test that is sensitive in detecting brain damage (23). The alternate finger-tapping task is also a psychomotor speed test, though one that requires less cognitive processing. In the absence of any impairment in the subjects’ attention and working memory task performance, we are unable to pinpoint the specific elements that would explain the psychomotor speed impairment among the post-COVID group.

### Increase in medical diagnoses and medical resource utilisation among post-COVID subjects

Post-COVID subjects had an increase in medical diagnoses since the COVID pandemic began, and showed an increase in hospitalisation in the past one year. This is consistent with other reports suggesting that there has been an increase in the incidence of some medical conditions after SARS-CoV-2 infection (24). The post-COVID group did not show an increase in post-COVID psychiatric diagnoses and new suicidal ideas, however. This suggests that, as a group, there is no evidence of clinically concerning mental health deterioration after infection, despite an increase in new diagnoses of physical conditions. The socioeconomic status of post-COVID subjects as compared to the matched control group did not worsen since the COVID pandemic began, and the HRQoL of both groups were comparable. Overall, these results reflect that, as a group, there has been some change in the health status of post-COVID subjects, even though this did not have significant impact on their social and vocational functioning and HRQoL.

### Post-COVID subjects with multiple neuropsychiatric complaints had poorer mental health and HRQoL

If we look at the comparison between those with a high load of persistent neuropsychiatric symptoms and those with a low load, however, the two groups have pervasively different post-COVID health trajectories despite similar pre-COVID health. This is clearly reflected by the new incidence of medical and psychiatric diagnoses, the emergence of suicidal ideation, increased medical consultations and hospitalisations and poor current mental health and worse current HRQoL of the post-COVID group, in the absence of significant pre-COVID health and socioeconomic discrepancies as compared to the matched control group. While we cannot tell the direction of any potential causal relationship between the neuropsychiatric and other aspects of health based on our data, given that this relationship may be bidirectional, or the two may share similar pathophysiology, the bottom-line is that the presence of multiple persistent post-COVID neuropsychiatric complaints reflected quantifiable and clinically relevant mental health distress. Notably, however, cognitive performance did not differ between the high and low symptom load groups. This can reflect the relationship between cognitive impairment and chronic post-COVID neuropsychiatric symptoms loading is non-linear. Yet, it could also have been a result of the limitations of the tools used. For example, because the cognitive tasks were completed via a mobile phone app in an uncontrolled environment, this may have increased the variance in performance.

### Lack of vaccination and pre-pandemic material deprivation predicts high number of chronic neuropsychiatric complaints but not index infection severity

Two of our three hypothesised risk factors, namely vaccination status before first COVID infection and pre-pandemic level of material deprivation as measured by the Deprivation Index, were found to be predictive factors of a high load of persistent neuropsychiatric symptoms. While previous studies have reported that COVID vaccinations reduce PACS (10), our study is the first to date that demonstrates that any COVID vaccination before infection reduces the risk of chronic neuropsychiatric symptoms even more than one year after infection, with this protective effect not being one that is mediated through reducing symptom severity or protection against reinfection. Thus, it is possible that vaccination reduces the dysregulated response of the immune system during SARS-CoV-2 infection, which is hypothesised to be the underlying pathophysiology of PACS. In this way, our findings highlight a further public health benefit of COVID vaccinations beyond reducing short-term morbidity and mortality.

Our results also suggest that socioeconomic factors cannot be overlooked when we are considering persistent post-COVID neuropsychiatric complaints. Material deprivation has been demonstrated to be a common contributing factor to mental health distress at the population level (Chung et al., 2021). In the context of the COVID pandemic, the association of poor COVID outcomes with health inequalities has been a recurrent theme(25,26). It is therefore not surprising that material deprivation was found to predict chronic post-COVID neuropsychiatric symptoms. While other studies have shown similar results, the limitations in the designs of such studies, namely in terms of the potential for reverse causality, have made it difficult to draw such conclusions straightforwardly from their results. A study from Hong Kong, for example, reported financial worry as one of the predictors of post-COVID mental health trajectory (27). Because they only measured the subjects’ evaluation of their financial situation post-COVID, however, there is difficulty determining the direction of causality given that a self-reported concern about financial status can be secondary to and a result of poor mental and physical health. A US-based study found that life stressors post-COVID are associated with neuropsychiatric outcomes 12 months post-infection (28), but the causal direction of this association is again unclear. To minimise concerns regarding reverse causality in our study, we collected pre-pandemic Deprivation Index data.

Contrary to our hypothesis, the severity of the index infection did not predict whether there would be a high or low load of chronic neuropsychiatric symptoms post-COVID. Available evidence on the ability of infection severity to predict PACS has not been consistent(27,29–32). There is convergent evidence, however, that persistent PACS can occur in individuals with mild infections.

### Embracing the heterogeneity among persistent neuropsychiatric complaints

Our network analysis suggested that the common chronic post-COVID neuropsychiatric symptoms can be grouped into two major (anxiety-depression and cognitive complaint-fatigue) and one minor (headache-dizziness) clusters, a finding which supports our hypothesis that such symptoms form clusters. These clusters suggest that there could be different sub-syndromes or even disorders under the umbrella term of persistent neuropsychiatric complaints.

We also found that socioeconomic disadvantage increased the symptom load of both major clusters, but that vaccination only significantly decreased the symptom load of the cognitive complaint-fatigue cluster. This difference in how predictive the factors were gives additional support to the idea that neuropsychiatric symptoms can be further categorised into subgroups, as different sub-syndromes may have different underlying risk factors and mechanisms. It is of interest that Chinese ethnicity positively predicts the number of symptoms in the cognitive complaint-fatigue cluster. Since there are only six non-ethnic Chinese subjects in the post-COVID group, however, this result is highly prone to selection bias and should be interpreted with caution.

Previous studies have also used clustering algorithms to categorise individuals with PACS into subgroups (33)(34). These algorithms usually use a mix of self-reported symptoms, assessment results, subject demographics and clinical factors to perform the clustering, whereas our approach is to first identify the clustering of symptoms before we examine the unique, associated factors of the discovered clusters. We argue that our approach is clearer phenomenologically because it reveals the relationship among symptoms independent of confounding sociodemographic factors. The key problem with any clustering result, however, is its generalisability. Although we have conducted extra bootstrapping procedures to demonstrate the stability of our clustering results, the further replication of these results using an independent cohort is important to support the generalisability of the clustering. To the best of our knowledge, Peter et al.’s study (Peter et al., 2022) is relatively comparable to ours in terms of study approach. Despite substantial differences in sampling, measurement and statistical tools employed, the results of their clustering analysis were largely consistent with ours: they found a cluster consisting of headache and dizziness, and another consisting of sleep, anxiety and depression.

## Conclusions

Our study suggests that the lack of COVID vaccination and a higher level of material deprivation before the COVID pandemic predicts a higher load of chronic post-COVID neuropsychiatric symptoms that persists for more than one year post-infection. This highlights the complex interplay of biological and socioeconomic factors that contribute to chronic post-COVID neuropsychiatric symptoms. Our analysis also suggests heterogeneity among chronic post-COVID neuropsychiatric symptoms, with two major symptom clusters (anxiety-depression and cognitive complaint-fatigue) and one minor cluster (headache-dizziness). Further study into chronic neuropsychiatric sequelae should take into account the heterogeneity and complexity of its phenomenology and aetiology.

## Strengths and limitations

The key strength of this study is that it was conducted in accordance with a pre-registered, open access protocol with pre-determined hypotheses. Another strength is that our assessment and analysis covered comprehensive demographic, clinical and socioeconomic factors, as well as symptoms and cognitive measures. The key limitation of this study is the use of the convenient sampling method, which limits the certainty of any epidemiological inference. Another major limitation is the cross-sectional design of the study. While we used the pre-COVID socioeconomic and health status of the subjects for case-control matching and regression analysis, so as to avoid bias introduced by changes in their physical and mental health due to COVID, the pre-COVID socioeconomic and health status data were based on recall, and thus may be affected by recall bias. The modest sample size also limits statistical power. Additionally, the infection status of the subjects was based on self-report and was not verified by our own testing, which could have rendered this data unreliable. Since Hong Kong implemented a ‘zero-COVID’ policy with very stringent surveillance prior to the Omicron wave in early 2022, however, the subjects’ understanding of their infection status is likely accurate. Lastly, we relied on self-reported checklists and symptom scales only, and did not conduct clinical interviews or use clinician-rated instruments.

## Data Availability

All data produced in the present study are available upon reasonable request to the authors.

## Acknowledgment

We thank Ms. Janet Tse for her logistical support in conducting the research. We thank Ms. Daisy Cheung for her very helpful comments and suggestions on the manuscript.

## Transparency Declaration

The lead author and manuscript guarantor (SWHC) affirm that the manuscript is an honest, accurate, and transparent account of the study being reported; that no important aspects of the study have been omitted; and that any discrepancies from the study as planned have been explained.

## Author contribution

SWHC, RNYC, PWCW, and WKW contributed to the design of the study. SWHC, RNYC, PWCW, SXL, YL, JWYC, PKSC, CKCL, TWHL, YKW contributed to the funding acquisition. SWHC, SXL, RNYC and YL contributed to in the design of assessment material. SWHC, TMC and YLL performed data analysis and wrote the first draft of the manuscript. All authors critically reviewed and approved the final manuscript.

## Data availability

The data that support the findings of this study will be openly available after the paper is published

## References

1. Nalbandian A, Sehgal K, Gupta A, Madhavan MV, McGroder C, Stevens JS, et al. Post-acute COVID-19 syndrome. Nat Med [Internet]. 2021 Apr [cited 2023 Sep 11];27(4):601–15. Available from: https://www.nature.com/articles/s41591-021-01283-z

2. White-Dzuro G, Gibson LE, Zazzeron L, White-Dzuro C, Sullivan Z, Diiorio DA, et al. Multisystem effects of COVID-19: a concise review for practitioners. Postgraduate Medicine [Internet]. 2021 Jan 2 [cited 2023 Aug 21];133(1):20–7. Available from: 10.1080/00325481.2020.1823094

3. Baillie JK, Lone NI, Jones S, Shaw A, Hairsine B, Kurasz C, et al. Multiorgan MRI findings after hospitalisation with COVID-19 in the UK (C-MORE): a prospective, multicentre, observational cohort study. The Lancet Respiratory Medicine [Internet]. 2023 Sep 22 [cited 2023 Sep 26];0(0). Available from: https://www.thelancet.com/journals/lanres/article/PIIS2213-2600(23)00262-X/fulltext

4. Davis HE, Assaf GS, McCorkell L, Wei H, Low RJ, Re’em Y, et al. Characterizing long COVID in an international cohort: 7 months of symptoms and their impact. EClinicalMedicine [Internet]. 2021 Aug 1 [cited 2021 Aug 27];38. Available from: https://www.thelancet.com/journals/eclinm/article/PIIS2589-5370(21)00299-6/abstract

5. Taquet M, Sillett R, Zhu L, Mendel J, Camplisson I, Dercon Q, et al. Neurological and psychiatric risk trajectories after SARS-CoV-2 infection: an analysis of 2-year retrospective cohort studies including 1 284 437 patients. The Lancet Psychiatry [Internet]. 2022 Oct 1 [cited 2023 Aug 14];9(10):815–27. Available from: https://www.thelancet.com/journals/lanpsy/article/PIIS2215-0366(22)00260-7/fulltext

6. Douaud G, Lee S, Alfaro-Almagro F, Arthofer C, Wang C, McCarthy P, et al. SARS-CoV-2 is associated with changes in brain structure in UK Biobank. Nature [Internet]. 2022 Apr [cited 2022 Jul 26];604(7907):697–707. Available from: https://www.nature.com/articles/s41586-022-04569-5

7. Besteher B, Machnik M, Troll M, Toepffer A, Zerekidze A, Rocktäschel T, et al. Larger gray matter volumes in neuropsychiatric long-COVID syndrome. Psychiatry Res [Internet]. 2022 Nov [cited 2023 Aug 24];317:114836. Available from: https://www.ncbi.nlm.nih.gov/pmc/articles/PMC9444315/

8. Besteher B, Rocktäschel T, Garza AP, Machnik M, Ballez J, Helbing DL, et al. Cortical thickness alterations and systemic inflammation define long-COVID patients with cognitive impairment [Internet]. Psychiatry and Clinical Psychology; 2023 Jul [cited 2023 Aug 24]. Available from: 10.1101/2023.07.21.23292988

9. Cosentino G, Todisco M, Hota N, Della Porta G, Morbini P, Tassorelli C, et al. Neuropathological findings from COVID-19 patients with neurological symptoms argue against a direct brain invasion of SARS-CoV-2: A critical systematic review. European Journal of Neurology [Internet]. 2021 [cited 2023 Aug 14];28(11):3856–65. Available from: 10.1111/ene.15045

10. Tsampasian V, Elghazaly H, Chattopadhyay R, Debski M, Naing TKP, Garg P, et al. Risk Factors Associated With Post-COVID-19 Condition: A Systematic Review and Meta-analysis. JAMA Intern Med. 2023 Jun 1;183(6):566–80.

11. Wong SC, Au AKW, Lo JYC, Ho PL, Hung IFN, To KKW, et al. Evolution and Control of COVID-19 Epidemic in Hong Kong. Viruses [Internet]. 2022 Nov 14 [cited 2023 Aug 10];14(11):2519. Available from: https://www.ncbi.nlm.nih.gov/pmc/articles/PMC9698160/

12. Chung RYN, Marmot M, Mak JKL, Gordon D, Chan D, Chung GKK, et al. Deprivation is associated with anxiety and stress. A population-based longitudinal household survey among Chinese adults in Hong Kong. J Epidemiol Community Health [Internet]. 2021 Apr 1 [cited 2023 Aug 23];75(4):335–42. Available from: https://jech.bmj.com/content/75/4/335

13. Saunders P, Wong H, Wong WP. Deprivation and Poverty in Hong Kong. Social Policy & Administration [Internet]. 2014 [cited 2023 Aug 23];48(5):556–75. Available from: https://onlinelibrary.wiley.com/doi/abs/10.1111/spol.12042

14. Wu KK, Chan KS. The development of the Chinese version of Impact of Event Scale--Revised (CIES-R). Soc Psychiatry Psychiatr Epidemiol. 2003 Feb;38(2):94–8.

15. Morin CM, Belleville G, Bélanger L, Ivers H. The Insomnia Severity Index: Psychometric Indicators to Detect Insomnia Cases and Evaluate Treatment Response. Sleep [Internet]. 2011 May 1 [cited 2023 Aug 23];34(5):601–8. Available from: https://www.ncbi.nlm.nih.gov/pmc/articles/PMC3079939/

16. Chalder T, Berelowitz G, Pawlikowska T, Watts L, Wessely S, Wright D, et al. Development of a fatigue scale. Journal of Psychosomatic Research [Internet]. 1993 Feb 1 [cited 2023 Aug 23];37(2):147–53. Available from: https://www.sciencedirect.com/science/article/pii/002239999390081P

17. Armstrong T, Bull F. Development of the World Health Organization Global Physical Activity Questionnaire (GPAQ). J Public Health [Internet]. 2006 Apr 1 [cited 2023 Aug 23];14(2):66–70. Available from: 10.1007/s10389-006-0024-x

18. The Whoqol Group. Development of the World Health Organization WHOQOL-BREF Quality of Life Assessment. Psychological Medicine [Internet]. 1998 May [cited 2023 Aug 23];28(3):551–8. Available from: https://www.cambridge.org/core/journals/psychological-medicine/article/abs/development-of-the-world-health-organization-whoqolbref-quality-of-life-assessment/0F50596B33A1ABD59A6605C44A6A8F30

19. Epskamp S, Borsboom D, Fried EI. Estimating psychological networks and their accuracy: A tutorial paper. Behav Res [Internet]. 2018 Feb 1 [cited 2023 Aug 18];50(1):195–212. Available from: 10.3758/s13428-017-0862-1

20. van Borkulo CD, Borsboom D, Epskamp S, Blanken TF, Boschloo L, Schoevers RA, et al. A new method for constructing networks from binary data. Sci Rep [Internet]. 2014 Aug 1 [cited 2023 Aug 18];4(1):5918. Available from: https://www.nature.com/articles/srep05918

21. Shizuka D, Farine DR. Measuring the robustness of network community structure using assortativity. Animal Behaviour [Internet]. 2016 Feb 1 [cited 2023 Aug 18];112:237–46. Available from: https://www.sciencedirect.com/science/article/pii/S0003347215004480

22. Peter RS, Nieters A, Kräusslich HG, Brockmann SO, Göpel S, Kindle G, et al. Post-acute sequelae of covid-19 six to 12 months after infection: population based study. BMJ [Internet]. 2022 Oct 13 [cited 2023 Aug 24];379:e071050. Available from: https://www.bmj.com/content/379/bmj-2022-071050

23. Jaeger J. Digit Symbol Substitution Test. J Clin Psychopharmacol [Internet]. 2018 Oct [cited 2023 Sep 22];38(5):513–9. Available from: https://www.ncbi.nlm.nih.gov/pmc/articles/PMC6291255/

24. Davis HE, McCorkell L, Vogel JM, Topol EJ. Long COVID: major findings, mechanisms and recommendations. Nat Rev Microbiol [Internet]. 2023 Mar [cited 2023 Sep 11];21(3):133–46. Available from: https://www.nature.com/articles/s41579-022-00846-2

25. Chung RYN, Chung GKK, Marmot M, Allen J, Chan D, Goldblatt P, et al. COVID-19 related health inequality exists even in a city where disease incidence is relatively low: a telephone survey in Hong Kong. J Epidemiol Community Health [Internet]. 2021 Jul [cited 2023 Oct 4];75(7):616–23. Available from: 10.1136/jech-2020-215392

26. Woodward M, Peters SAE, Harris K. Social deprivation as a risk factor for COVID-19 mortality among women and men in the UK Biobank: nature of risk and context suggests that social interventions are essential to mitigate the effects of future pandemics. J Epidemiol Community Health [Internet]. 2021 Nov [cited 2023 Oct 4];75(11):1050–5. Available from: 10.1136/jech-2020-215810

27. Leung ONW, Chiu NKH, Wong SYS, Cuijpers P, Alonso J, Chan PKS, et al. Dimensional structure of one-year post-COVID-19 neuropsychiatric and somatic sequelae and association with role impairment. Sci Rep [Internet]. 2023 Jul 27 [cited 2023 Aug 2];13(1):12205. Available from: https://www.nature.com/articles/s41598-023-39209-z

28. Frontera JA, Sabadia S, Yang D, de Havenon A, Yaghi S, Lewis A, et al. Life stressors significantly impact long-term outcomes and post-acute symptoms 12-months after COVID-19 hospitalization. J Neurol Sci [Internet]. 2022 Dec 15 [cited 2023 Aug 24];443:120487. Available from: https://www.ncbi.nlm.nih.gov/pmc/articles/PMC9637014/

29. Ceban F, Ling S, Lui LMW, Lee Y, Gill H, Teopiz KM, et al. Fatigue and cognitive impairment in Post-COVID-19 Syndrome: A systematic review and meta-analysis. Brain, Behavior, and Immunity [Internet]. 2022 Mar 1 [cited 2023 Aug 2];101:93–135. Available from: https://www.sciencedirect.com/science/article/pii/S0889159121006516

30. Elmazny A, Magdy R, Hussein M, Elsebaie EH, Ali SH, Abdel Fattah AM, et al. Neuropsychiatric post-acute sequelae of COVID-19: prevalence, severity, and impact of vaccination. Eur Arch Psychiatry Clin Neurosci [Internet]. 2023 Jan 27 [cited 2023 Aug 21]; Available from: 10.1007/s00406-023-01557-2

31. Evans RA, McAuley H, Harrison EM, Shikotra A, Singapuri A, Sereno M, et al. Physical, cognitive, and mental health impacts of COVID-19 after hospitalisation (PHOSP-COVID): a UK multicentre, prospective cohort study. The Lancet Respiratory Medicine [Internet]. 2021 Nov [cited 2022 Oct 31];9(11):1275–87. Available from: https://linkinghub.elsevier.com/retrieve/pii/S2213260021003830

32. Townsend L, Dyer AH, Jones K, Dunne J, Mooney A, Gaffney F, et al. Persistent fatigue following SARS-CoV-2 infection is common and independent of severity of initial infection. PLOS ONE [Internet]. 2020 Nov 9 [cited 2023 Aug 2];15(11):e0240784. Available from: https://journals.plos.org/plosone/article?id=10.1371/journal.pone.0240784

33. Evans RA, McAuley H, Harrison EM, Shikotra A, Singapuri A, Sereno M, et al. Physical, cognitive, and mental health impacts of COVID-19 after hospitalisation (PHOSP-COVID): a UK multicentre, prospective cohort study. The Lancet Respiratory Medicine [Internet]. 2021 Nov 1 [cited 2023 Aug 17];9(11):1275–87. Available from: https://www.thelancet.com/journals/lanres/article/PIIS2213-2600(21)00383-0/fulltext

34. Zhao Y, Shi L, Jiang Z, Zeng N, Mei H, Lu Y, et al. The phenotype and prediction of long-term physical, mental and cognitive COVID-19 sequelae 20 months after recovery, a community-based cohort study in China. Mol Psychiatry [Internet]. 2023 Jan 23 [cited 2023 Apr 10];1–9. Available from: https://www.ncbi.nlm.nih.gov/pmc/articles/PMC9869317/

