## Supplementary materials for "Chronic post-COVID neuropsychiatric symptoms persisting beyond one year from infection: a case-control study and network analysis"

*Full Methodology*

To explore relationships between chronic long-COVID symptoms and their clustering patterns, we built a **regularized partial correlation network** based on the self-report symptoms data: each node (variable) represented a chronic long-COVID symptom (such as fatigue, headache, daytime sleepiness); each edge between nodes represented the regularized **partial** correlation between the symptoms, controlling for the effects of other symptoms in the network model (Hevey, 2018). To minimize the instability of the network, if there were only fewer than or equal to 20 people (out of a sample of 223 people) who suffered from a particular symptom, the node representing that symptom would be removed from the network model at the outset. As the model fully consisted of binary variables, an *Ising model* was used to estimate the network from the symptoms data, with LASSO regularization that reduced spurious partial correlations / edges and resulted in a more parsimonious model (Friedman et al., 2008; Hevey, 2018; van Borkulo et al., 2014). When estimating the network model, a number of network models was being estimated under different values of the LASSO's tuning parameter lambda. Subsequently, we selected the model with the lowest Extended Bayesian Information Criterion (Chen & Chen, 2008). R package *bootnet* and *IsingFIt* were used to complete the specified network estimation process (Epskamp et al., 2018; van Borkulo et al., 2014). Additional R packages were used: *igraph* (Csardi & Nepusz, 2006), *qgraph* (Epskamp et al., 2012) and *plyr* (Wickham, 2011). After network estimation, we used a community detection algorithm, *walktrap algorithm* to find out the communities / clusters within the symptom network (Pons & Latapy, 2005). As the *walktrap algorithm* cannot handle negative edge values, we reset those edges to a value of 0 (Gates et al., 2016).

<u>Validation of Community Detection</u>

To validate the robustness of the detected communities, we used a metric called *community assortativity* ($R_{com}$) to measure the robustness of community assignment done by the *walktrap algorithm*. To calculate *community assortativity*, we first generated 1000 bootstrap replicate networks (networks estimated from re-sampling the symptoms data **with** replacement); among these 1000 replicate networks, we measured "the degree to which pairs assigned to the same community in the empirical network also occur in the same community in bootstrap replicate networks" (Shizuka & Farine, 2016). If a network's $R_{com}$ is equal to 1, it implies the highest robustness of community assignment, where community assignments in all bootstrap replicate networks were identical to those in the empirical network. This also indicates that the network likely contains discrete clusters. If $R_{com}$ is equal to 0, it implies that community assignments were essentially random with regard to the original community assignments in the empirical network. The community assignments are deemed to be robust if $R_{com}$ is larger than 0.5 (Shizuka & Farine, 2016).

The $R_{com}$ of the network generated based on our symptoms data was 0.68, indicating our network model likely contained discrete clusters and the *walktrap algorithm* reliably detected those clusters / communities. *Community assortativity* was calculated using adapting code provided by (Shizuka & Farine, 2016). Additional R packages were used in this section: *asnipe* (Farine, 2013) and *assortnet* (Farine, 2014).

*List of Post-acute COVID syndrome (PACS) Checklist*

| | |
|---|---|
| **15 neuropsychiatric items** | |
| | Memory problems |
| | Fatigue |
| | Inability to concentrate |
| | Feeling anxious |
| | Insomnia |
| | Daytime Sleepiness |
| | Feeling depressed |
| | Headache |
| | Loss of interest or pleasure |
| | Dizziness |
| | Tinnitus |
| | Loss or change to your sense of taste and smell |
| | auditory hallucinations, delirium symptoms |
| | Move with difficulty |
| | Loss of hearing |
| **26 non-neuropsychiatric items** | |
| | Loss of hair |
| | Shortness of breath or trouble breathing |
| | Generalised discomfort |
| | Low back and/or back pain |
| | Body weaknesses |
| | Joint pain |
| | Cough |
| | Chest congestion and /or Chest pain |
| | Muscle pain |
| | Heart palpitations and /or irregular heartbeat |
| | Blurred Vision |
| | Night sweats |
| | Congested or stuffy nose |
| | Abdominal pain and/or bloating and/or heartburn |
| | Constipation |
| | Diarrhoea and/or nausea or vomiting |
| | Sore or painful throat |
| | Runny or dripping nose |
| | Rashes |
| | Sneeze |
| | Fever/High temperature |
| | Loss of appetite |
| | Dysmenorrhea |
| | Conjunctivitis |
| | Earaches and/or ear numbness |
| | Red, swollen and/or strange patches on the tongue |

*Note.* Although we include post-traumatic stress (PTS) in our network analysis and symptom clustering as one of the neuropsychiatric symptoms, this symptom is measured using the Impact of Event Scale-Revised (IES-R) questionnaire, but it is not included in the original PACS symptom checklist.

*Cognitive Test Instructions*

**Reaction Test**

1: Please pay attention to the black box at the center of the screen.

2: When a red number appears in the box, click anywhere on the screen as soon as possible.

3: Avoid clicking on the screen before the numbers appear.

**Number Symbol Test**

1: A symbol will appear in the center of the screen.

2: Please find the number corresponding to this symbol from the blue table at the top of the screen.

3: And click the orange button with the corresponding number at the bottom of the screen.

**Memory Test**

**1-Back memory test**

1: In this test, different English letters will be presented in sequence. If the current letter presented is identical to the previous (1) letter, click “Yes”, otherwise click “No”. Please do this as quickly and accurately as possible.

2: In this test, a sequence of letters will be presented one after another. When the current letter is presented, you need to judge whether it is the same as the previous (1) letter, if it is the same, click "Yes"; if it is different, click “No”. In the following example, [M, H, F, F] will be presented one by one in order. You need to click “No” when the first three letters [M, H, F] are presented. And when you see the fourth letter [F], you need to click “Yes” because this letter is the same as the previous (1) letter.

**2-Back memory test**

1: In this test, different English letters will be presented in sequence. If the current letter presented is identical to the previous (2) letters, click “Yes”, otherwise click “No”. Please do this as quickly and accurately as possible.

2: In this test, a sequence of letters will be presented one after another. When the current letter is presented, you need to judge whether it is the same as the previous (2) letters, if it is the same, click “Yes”; if it is different, click “No”. In the following example, [M, H, F, F] will be presented one by one in order. You need to click “No” when the first three letters [M, H, F] are presented. And when you see the fourth letter [F], you need to click “Yes” because this letter is the same as the previous (2) letters.

**Finger Reaction and Coordination Assessment**

1: Please use your index and middle fingers to click the pink button on the screen alternately within 10-seconds test time. Each pink button click scores 1 point.

Please click with your index finger first (1)

Click again with your middle finger (2)

*Participant Recruitment Flowcharts for COVID group and Control group, respectively*

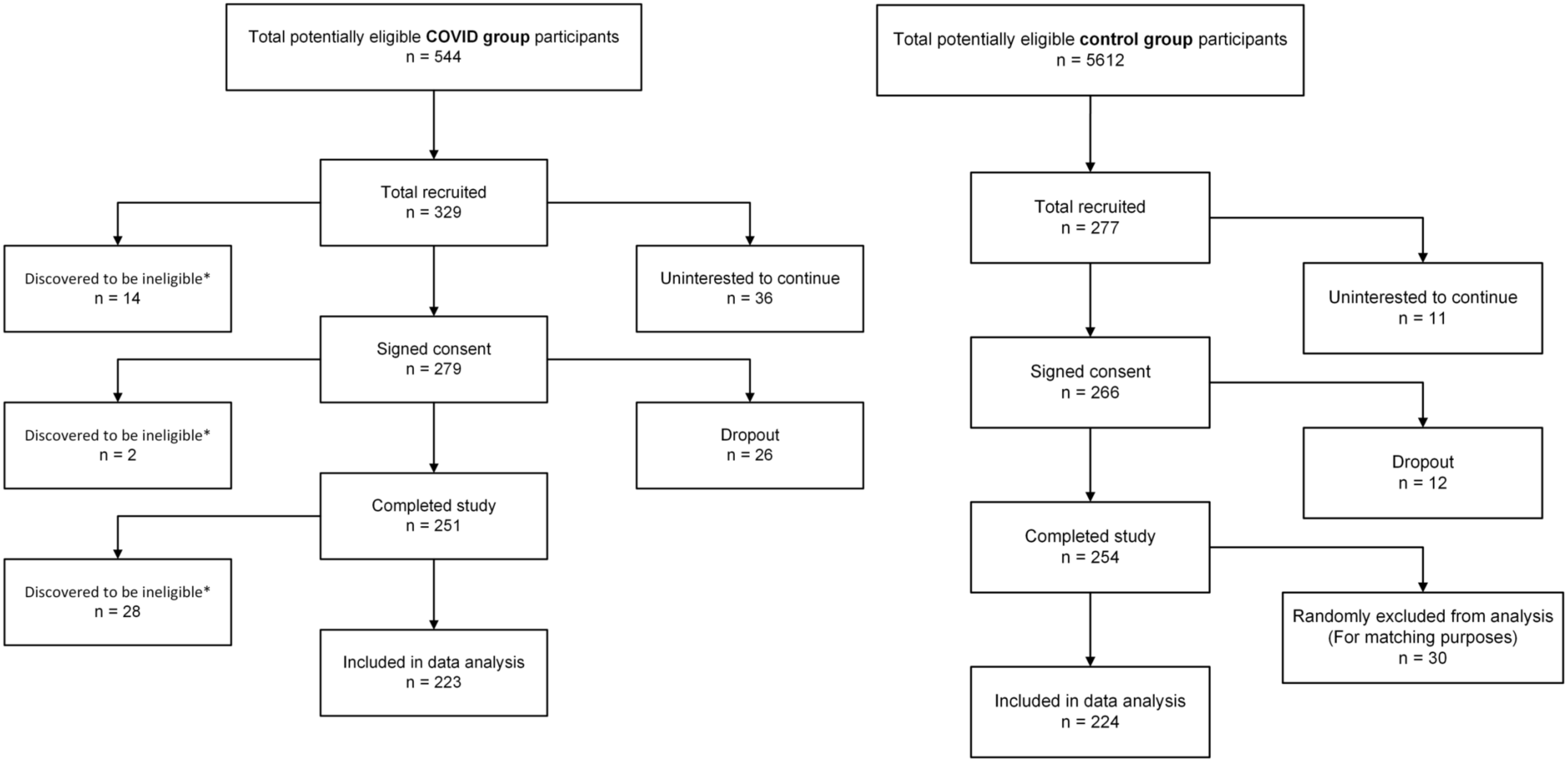

*Relationship between the number of chronic neuropsychiatric symptoms (x-axis) and mental health measurements (y-axis)*

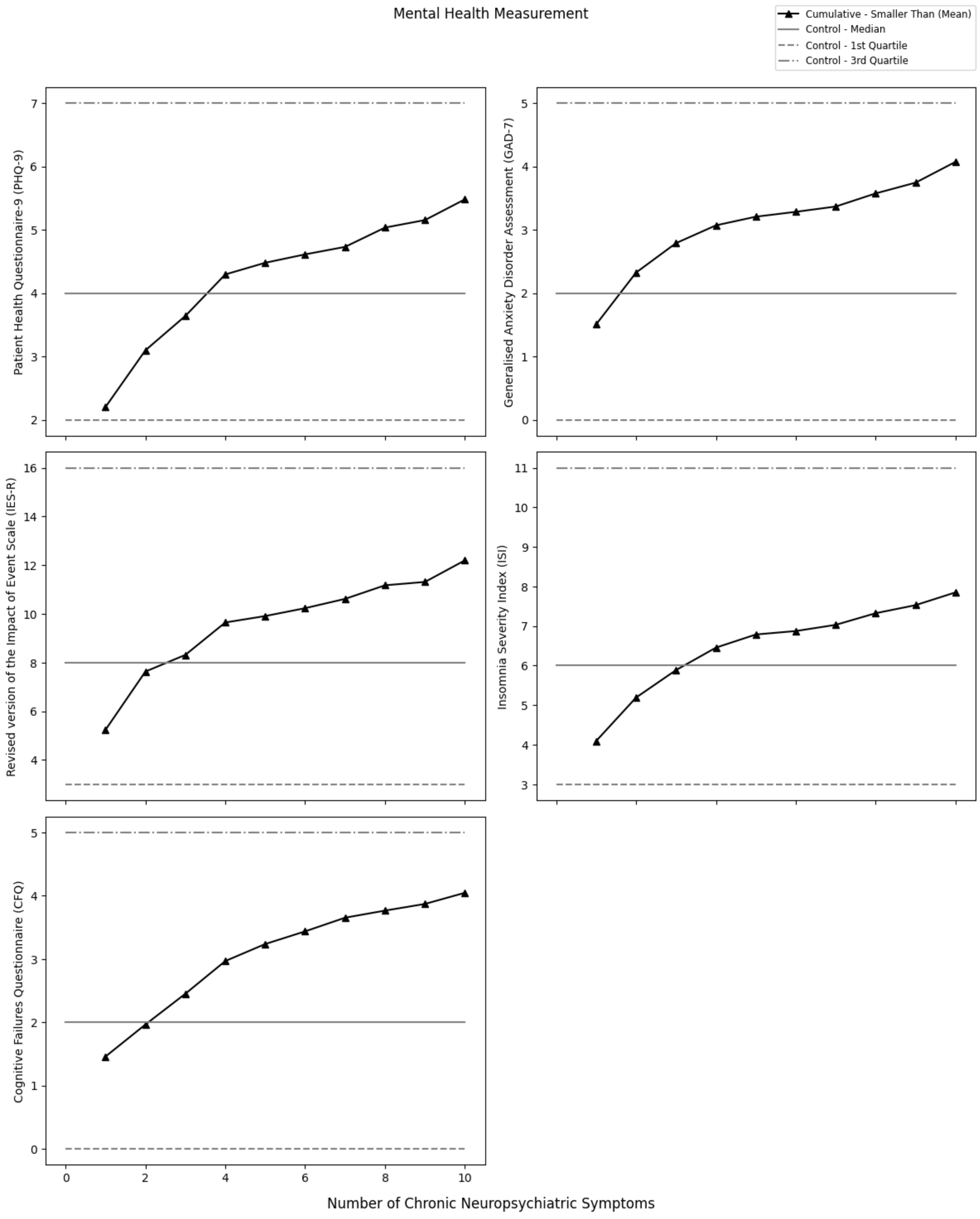

*Relationship between the number of chronic neuropsychiatric symptoms (x-axis) and cognitive task performance (y-axis)*

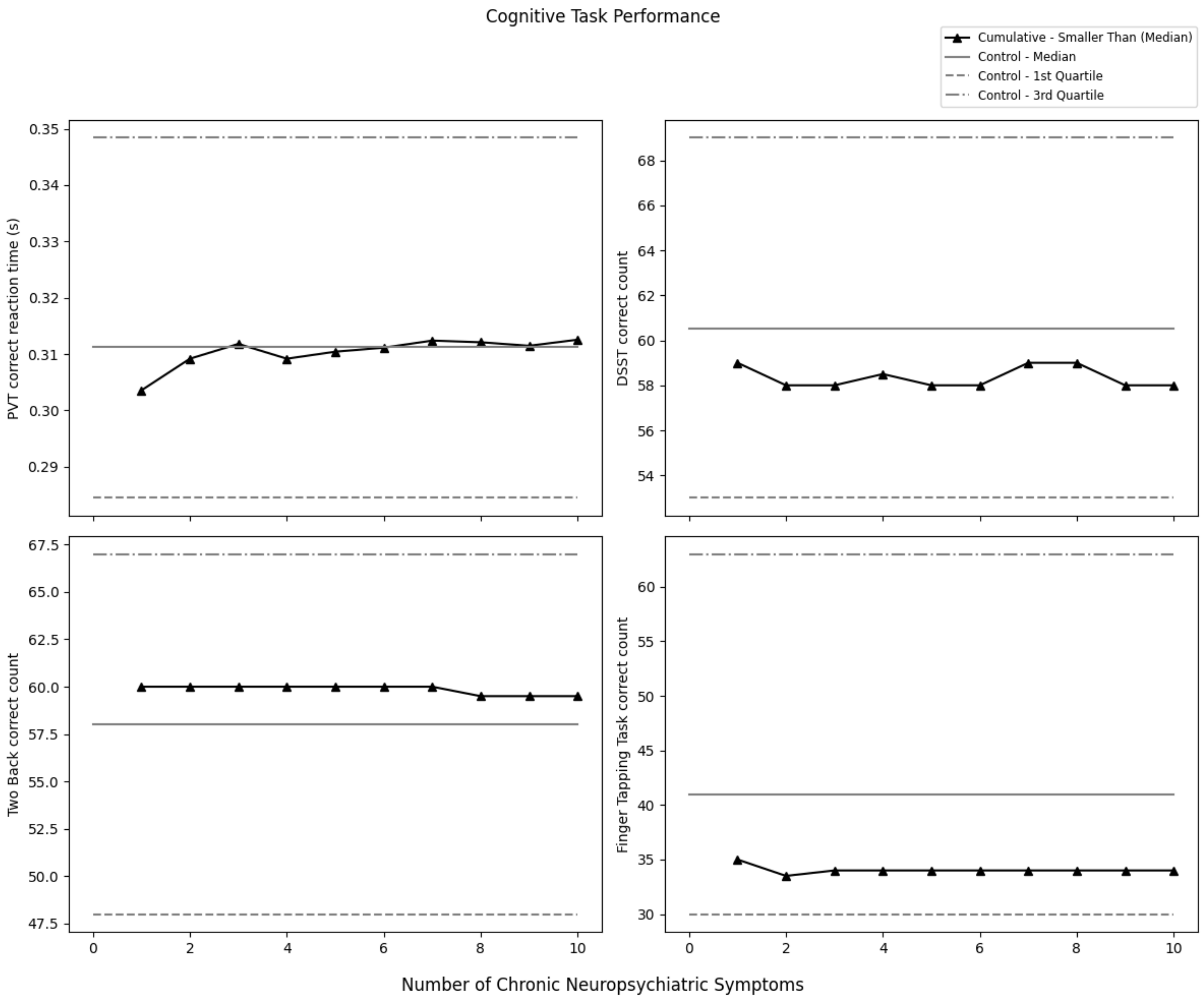

*Relationship between the number of chronic neuropsychiatric symptoms (x-axis) and health-related quality of life outcome (y-axis)*

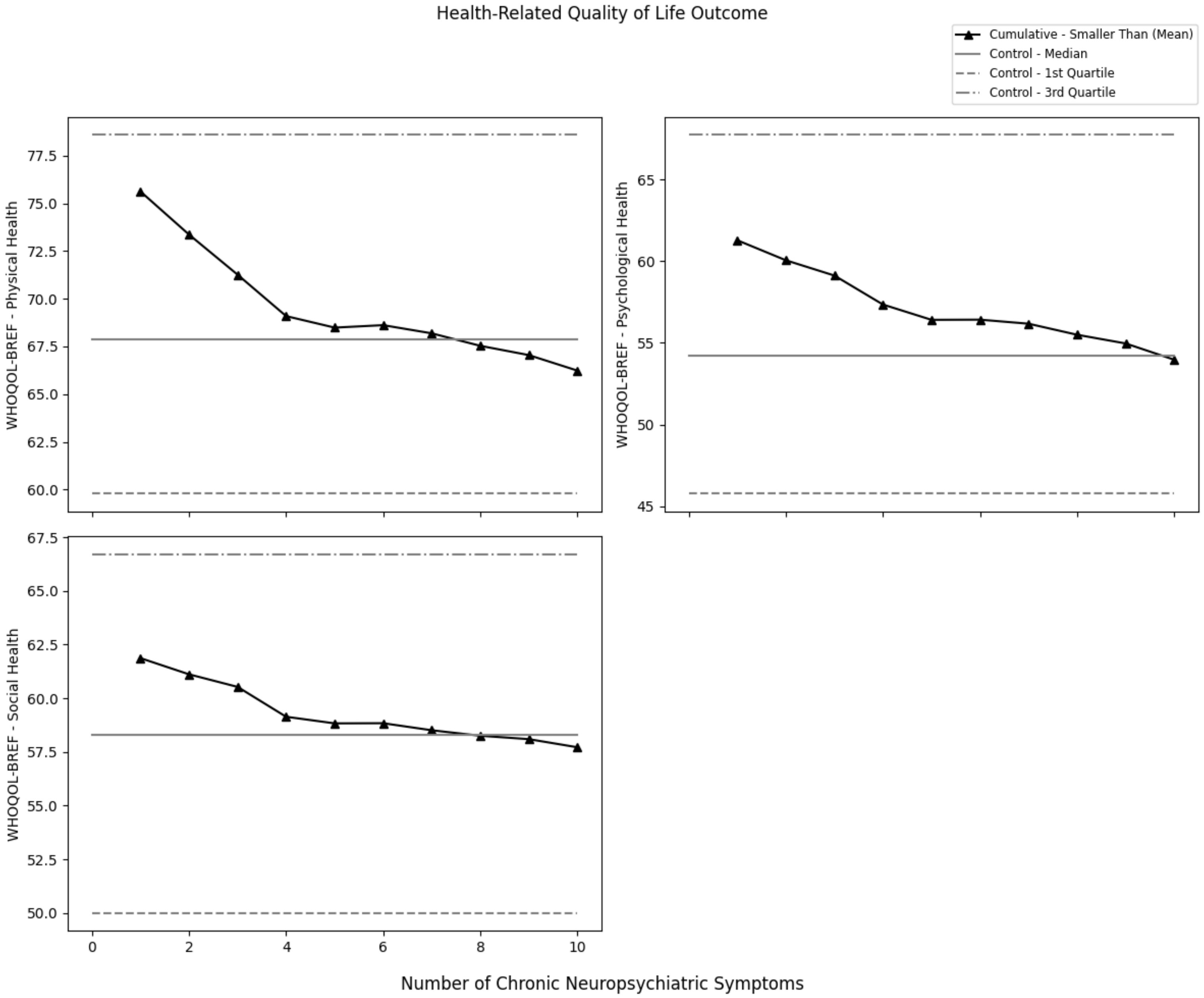

*Comparison of **pre-pandemic** medical diagnoses between subjects of post-COVID group and control group, and between high post-COVID neuropsychiatric symptom load group and low symptom load group*

| Medical Diagnoses | COVID Group n (%) | Control Group n (%) | High Symptoms Load Group n (%) | Low Symptoms Load Group n (%) |
|---|---|---|---|---|
| None | 180.0 (80.7) | 180.0 (80.4) | 51.0 (76.1) | 309.0 (81.3) |
| Hypertension | 18.0 (8.1) | 22.0 (9.8) | 7.0 (10.4) | 33.0 (8.7) |
| Chronic (long-term) respiratory diseases, such as asthma, chronic obstructive pulmonary disease (COPD), emphysema or bronchitis | 4.0 (1.8) | 3.0 (1.3) | 1.0 (1.5) | 6.0 (1.6) |
| Chronic heart disease such as heart failure | 5.0 (2.2) | 2.0 (0.9) | 2.0 (3.0) | 5.0 (1.3) |
| Chronic kidney disease | 0.0 (0.0) | 0.0 (0.0) | 0.0 (0.0) | 0.0 (0.0) |
| Chronic liver disease, such as hepatitis | 6.0 (2.7) | 5.0 (2.2) | 3.0 (4.5) | 8.0 (2.1) |
| Chronic gastrointestinal disease, such as peptic ulcer | 2.0 (0.9) | 3.0 (1.3) | 1.0 (1.5) | 4.0 (1.1) |
| Major or minor stroke | 2.0 (0.9) | 0.0 (0.0) | 1.0 (1.5) | 1.0 (0.3) |
| Chronic neurological conditions (except stroke), such as Parkinson's disease, motor neuron disease, multiple sclerosis (MS), a learning disability or cerebral palsy | 1.0 (0.4) | 1.0 (0.4) | 0.0 (0.0) | 2.0 (0.5) |
| Diabetes | 13.0 (5.8) | 7.0 (3.1) | 1.0 (1.5) | 19.0 (5.0) |
| A weakened immune system as the result of conditions such as HIV and AIDS, or immune-suppressing medicines such as steroid tablets or chemotherapy | 1.0 (0.4) | 1.0 (0.4) | 0.0 (0.0) | 2.0 (0.5) |
| Being seriously overweight (a body mass index (BMI) of 30 or above) | 0.0 (0.0) | 2.0 (0.9) | 0.0 (0.0) | 2.0 (0.5) |
| Organ transplant recipient | 0.0 (0.0) | 1.0 (0.4) | 0.0 (0.0) | 1.0 (0.3) |
| Cancer, receiving treatment | 0.0 (0.0) | 0.0 (0.0) | 0.0 (0.0) | 0.0 (0.0) |
| Cancer, completed treatment | 2.0 (0.9) | 1.0 (0.4) | 0.0 (0.0) | 3.0 (0.8) |
| Chronic pain | 7.0 (3.1) | 14.0 (6.2) | 4.0 (6.0) | 17.0 (4.5) |

*Comparison of **newly diagnosed** medical diagnoses since the pandemic began between subjects of COVID group and control group, and between high post-COVID neuropsychiatric symptom load group and low symptom load group*

| Medical Diagnoses | COVID Group n (%) | Control Group n (%) | High Symptoms Load Group n (%) | Low Symptoms Load Group n (%) |
|---|---|---|---|---|
| None | 196.0 (87.9) | 213.0 (95.1) | 52.0 (77.6) | 357.0 (93.9) |
| Hypertension | 9.0 (4.0) | 5.0 (2.2) | 5.0 (7.5) | 9.0 (2.4) |
| Chronic (long-term) respiratory diseases, such as asthma, chronic obstructive pulmonary disease (COPD), emphysema or bronchitis | 2.0 (0.9) | 0.0 (0.0) | 0.0 (0.0) | 2.0 (0.5) |
| Chronic heart disease such as heart failure | 3.0 (1.3) | 1.0 (0.4) | 3.0 (4.5) | 1.0 (0.3) |
| Chronic kidney disease | 0.0 (0.0) | 0.0 (0.0) | 0.0 (0.0) | 0.0 (0.0) |
| Chronic liver disease, such as hepatitis | 2.0 (0.9) | 1.0 (0.4) | 0.0 (0.0) | 3.0 (0.8) |
| Chronic gastrointestinal disease, such as peptic ulcer | 0.0 (0.0) | 0.0 (0.0) | 0.0 (0.0) | 0.0 (0.0) |
| Major or minor stroke | 0.0 (0.0) | 0.0 (0.0) | 0.0 (0.0) | 0.0 (0.0) |
| Chronic neurological conditions (except stroke), such as Parkinson’s disease, motor neuron disease, multiple sclerosis (MS), a learning disability or cerebral palsy | 0.0 (0.0) | 0.0 (0.0) | 0.0 (0.0) | 0.0 (0.0) |
| Diabetes | 6.0 (2.7) | 0.0 (0.0) | 2.0 (3.0) | 4.0 (1.1) |
| A weakened immune system as the result of conditions such as HIV and AIDS, or immune-suppressing medicines such as steroid tablets or chemotherapy | 0.0 (0.0) | 0.0 (0.0) | 0.0 (0.0) | 0.0 (0.0) |
| Being seriously overweight (a body mass index (BMI) of 30 or above) | 1.0 (0.4) | 0.0 (0.0) | 0.0 (0.0) | 1.0 (0.3) |
| Organ transplant recipient | 0.0 (0.0) | 0.0 (0.0) | 0.0 (0.0) | 0.0 (0.0) |
| Cancer, receiving treatment | 0.0 (0.0) | 0.0 (0.0) | 0.0 (0.0) | 0.0 (0.0) |
| Cancer, completed treatment | 0.0 (0.0) | 2.0 (0.9) | 0.0 (0.0) | 2.0 (0.5) |
| Chronic pain | 9.0 (4.0) | 3.0 (1.3) | 7.0 (10.4) | 5.0 (1.3) |

*Comparison of **pre-pandemic** psychiatric diagnoses between subjects of COVID group and control group, and between high post-COVID neuropsychiatric symptom load group and low symptom load group*

| Psychiatric Diagnoses | COVID Group<br>n (%) | Control Group<br>n (%) | High Symptoms Load Group<br>n (%) | Low Symptoms Load Group<br>n (%) |
|---|---|---|---|---|
| None | 212.0 (95.1) | 213.0 (95.1) | 64.0 (95.5) | 361.0 (95.0) |
| Substance use disorder | 0.0 (0.0) | 0.0 (0.0) | 0.0 (0.0) | 0.0 (0.0) |
| Bipolar disorder | 2.0 (0.9) | 2.0 (0.9) | 0.0 (0.0) | 4.0 (1.1) |
| Obsessive-compulsive disorder | 0.0 (0.0) | 0.0 (0.0) | 0.0 (0.0) | 0.0 (0.0) |
| Social phobia | 2.0 (0.9) | 0.0 (0.0) | 2.0 (3.0) | 0.0 (0.0) |
| Other anxiety disorder | 3.0 (1.3) | 1.0 (0.4) | 2.0 (3.0) | 2.0 (0.5) |
| Psychotic disorder | 0.0 (0.0) | 0.0 (0.0) | 0.0 (0.0) | 0.0 (0.0) |
| Depression | 3.0 (1.3) | 7.0 (3.1) | 0.0 (0.0) | 10.0 (2.6) |
| Generalised anxiety disorder | 3.0 (1.3) | 3.0 (1.3) | 1.0 (1.5) | 5.0 (1.3) |
| Post-traumatic stress disorder | 0.0 (0.0) | 0.0 (0.0) | 0.0 (0.0) | 0.0 (0.0) |
| Panic disorder | 1.0 (0.4) | 0.0 (0.0) | 0.0 (0.0) | 1.0 (0.3) |
| Eating disorder | 1.0 (0.4) | 0.0 (0.0) | 0.0 (0.0) | 1.0 (0.3) |

*Comparison of **newly diagnosed** psychiatric diagnoses since the pandemic began between subjects of COVID group and control group, and between high post-COVID neuropsychiatric symptoms load group and low symptoms load group*

| Psychiatric Diagnoses | COVID Group n (%) | Control Group n (%) | High Symptoms Load Group n (%) | Low Symptoms Load Group n (%) |
|---|---|---|---|---|
| None | 217.0 (97.3) | 217.0 (96.9) | 62.0 (92.5) | 372.0 (97.9) |
| Substance use disorder | 0.0 (0.0) | 0.0 (0.0) | 0.0 (0.0) | 0.0 (0.0) |
| Bipolar disorder | 0.0 (0.0) | 1.0 (0.4) | 0.0 (0.0) | 1.0 (0.3) |
| Obsessive-compulsive disorder | 0.0 (0.0) | 0.0 (0.0) | 0.0 (0.0) | 0.0 (0.0) |
| Social phobia | 0.0 (0.0) | 1.0 (0.4) | 0.0 (0.0) | 1.0 (0.3) |
| Other anxiety disorder | 2.0 (0.9) | 1.0 (0.4) | 2.0 (3.0) | 1.0 (0.3) |
| Psychotic disorder | 0.0 (0.0) | 0.0 (0.0) | 0.0 (0.0) | 0.0 (0.0) |
| Depression | 2.0 (0.9) | 5.0 (2.2) | 2.0 (3.0) | 5.0 (1.3) |
| Generalised anxiety disorder | 0.0 (0.0) | 2.0 (0.9) | 0.0 (0.0) | 2.0 (0.5) |
| Post-traumatic stress disorder | 1.0 (0.4) | 1.0 (0.4) | 1.0 (1.5) | 1.0 (0.3) |
| Panic disorder | 0.0 (0.0) | 2.0 (0.9) | 0.0 (0.0) | 2.0 (0.5) |
| Eating disorder | 1.0 (0.4) | 1.0 (0.4) | 0.0 (0.0) | 2.0 (0.5) |

*Logistic regression model predicting symptom load (high / low)*

| Variables | Coefficient | SE | *p* | Coef CI [5%] | Coef CI [95%] | AOR | AOR [5%] | AOR [95%] |
|---|---|---|---|---|---|---|---|---|
| Intercept | -1.63 | 0.78 | 0.037 | -2.91 | -0.34 | 0.20 | 0.04 | 0.91 |
| Age | 0.02 | 0.01 | 0.23 | -0.01 | 0.04 | 1.02 | 0.99 | 1.04 |
| Gender | -0.01 | 0.29 | 0.98 | -0.49 | 0.48 | 0.99 | 0.56 | 1.77 |
| Non-ethnic Chinese | -14.07 | 630.41 | 0.98 | -1051.00 | 1022.85 | 0.00 | 0.00 | infinity |
| Tertiary education | 0.06 | 0.36 | 0.87 | -0.53 | 0.65 | 1.06 | 0.52 | 2.14 |
| Pre-covid deprivation index (log-transformed) | 0.44 | 0.21 | 0.033 | 0.10 | 0.79 | 1.56 | 1.04 | 2.35 |
| Pre-covid no. of medical conditions | -0.08 | 0.26 | 0.75 | -0.50 | 0.34 | 0.92 | 0.56 | 1.53 |
| Pre-covid no. of psychiatric conditions | 0.11 | 0.76 | 0.88 | -1.14 | 1.36 | 1.12 | 0.25 | 4.96 |
| Severity of first covid infection | 0.02 | 0.22 | 0.91 | -0.34 | 0.39 | 1.03 | 0.67 | 1.58 |
| More than 1 covid infection | -0.35 | 0.36 | 0.32 | -0.94 | 0.23 | 0.70 | 0.35 | 1.41 |
| Received vaccine(s) before first covid infection | -1.37 | 0.66 | 0.036 | -2.45 | -0.29 | 0.25 | 0.07 | 0.92 |

*Note.* AOR: adjusted odds ratio.

*Linear regression model predicting the number of symptoms from the anxiety-depression cluster*

| Variables | Unstandardized Coefficients | | Standardized Coefficients | | |
|---|---|---|---|---|---|
| | B | Std. Error | Beta | t | Sig. |
| | 0.715 | 0.464 | | 1.541 | 0.125 |
| Age | 0.005 | 0.008 | 0.052 | 0.632 | 0.528 |
| Gender | -0.104 | 0.173 | -0.042 | -0.600 | 0.549 |
| Non-ethnic Chinese | -0.807 | 0.582 | -0.096 | -1.387 | 0.167 |
| Tertiary education | -0.042 | 0.218 | -0.015 | -0.191 | 0.848 |
| Pre-covid deprivation index (log-transformed) | 0.319 | 0.128 | 0.177 | 2.492 | 0.013 |
| Pre-covid no. of medical conditions | -0.085 | 0.158 | -0.039 | -0.535 | 0.593 |
| Pre-covid no. of psychiatric conditions | 0.509 | 0.458 | 0.077 | 1.111 | 0.268 |
| Severity of first covid infection | -0.111 | 0.139 | -0.056 | -0.798 | 0.426 |
| More than 1 covid infection | -0.268 | 0.211 | -0.086 | -1.270 | 0.206 |
| Received vaccine(s) before first covid infection | -0.434 | 0.300 | -0.100 | -1.450 | 0.149 |

*Linear regression model predicting the number of symptoms from the cognitive complaints-fatigue cluster*

| Variables | Unstandardized Coefficients | | Standardized Coefficients | | |
|---|---|---|---|---|---|
| | B | Std. Error | Beta | t | Sig. |
| Intercept | 1.157 | 0.484 | | 2.393 | 0.018 |
| Age | 0.000 | 0.008 | 0.003 | 0.038 | 0.969 |
| Gender | 0.050 | 0.181 | 0.019 | 0.279 | 0.781 |
| Non-ethnic Chinese | -1.264 | 0.607 | -0.143 | -2.083 | 0.038 |
| Tertiary education | -0.076 | 0.228 | -0.026 | -0.332 | 0.740 |
| Pre-covid deprivation index (log-transformed) | 0.293 | 0.133 | 0.155 | 2.197 | 0.029 |
| Pre-covid no. of medical conditions | -0.004 | 0.165 | -0.002 | -0.025 | 0.980 |
| Pre-covid no. of psychiatric conditions | 0.521 | 0.478 | 0.075 | 1.091 | 0.277 |
| Severity of first covid infection | 0.036 | 0.145 | 0.017 | 0.248 | 0.805 |
| More than 1 covid infection | -0.004 | 0.220 | -0.001 | -0.018 | 0.985 |
| Received vaccine(s) before first covid infection | -0.715 | 0.312 | -0.158 | -2.289 | 0.023 |
